# A multimodal exploration of circulating inflammatory markers in patients undergoing surgical intervention for lumbar disc herniation in selected hospitals of Sri Lanka

**DOI:** 10.64898/2026.04.21.26351426

**Authors:** Piriyanka Aravinth, Niroshima Dedunu Withanage, Bhagya Madhubhanie Senadheera, Sujatha Pathirge, Sinnathamby Ponnampalam Athiththan, Sunil Lakshman Perera, Lohini Vijayendran Athiththan

## Abstract

**Background:** Inflammatory markers play an important role in the pathophysiology of Lumbar disc herniation (LDH). This study presents a comprehensive multi-assessment of the inflammatory landscape by combining serum inflammatory cytokines quantification, their diagnostic performance, associations with radiological features, and integrating the experimental findings into an in-silico protein–protein interaction network.

**Methods:** A multifaceted study design was utilized to quantify and compare the distribution of selected inflammatory cytokines in patients with LDH and control subjects. The diagnostic ability of these cytokines was assessed using receiver operating characteristic curve analysis. The cytokines values were correlated with selected radiological findings including disc herniation subtypes (protrusion, extrusion, and sequestration), and further categorized as contained and non-contained in patients using a Spearman’s rank correlation test. Additionally, computational analysis was performed to identify the central hubs and functionally enriched pathways.

**Results:** In patients with LDH, IL-6 and IL-1β showed statistically significant (IL-6: p < 0.001; IL-1β: p = 0.001) rise, but IL-6 showed high diagnostic and discriminative power (AUC = 0.99; cut-off: 19.99 pg/mL). Further IL-1β exhibited a positive correlation with non-contained disc herniation (extrusion and sequestration), while displaying a significant (p < 0.05) negative correlation with protrusion. In silico analysis identified IL-1β, IL-8, TNF-α, IL-6, IL-1α, CSF2, CSF3, and IL-10 as central hubs, with IL-1β being the top ranked hub in determining functionally enriched cytokine-cytokine receptor interaction.

**Conclusions:** Study confirmed IL-6 as a powerful diagnostic marker for LDH, while IL-1β aids in determining contained and non-contained disc herniation. Further, IL-1β was identified as the central hub, triggering functionally enriched pathways in the pathogenesis of LDH.

## Introduction

Globally, low back pain (LBP) is a major contributing factor for years lived with disability where half a billion cases were recorded in the year 2020 [1].

Lumbar disc herniation (LDH) often leads to LBP, lumbar radicular pain [2], lumbar radiculopathy [3], neurogenic claudication, and cauda equina syndrome [4,5]. LDH is closely related with persistent LBP, with affected patients showing dramatically higher prevalence, compared to the general population (46.2% vs 11.9%) [6].

Traditionally, mechanical compression of nerve roots caused by the herniated discs has been considered as the major contributing factor associated with the worsening signs and symptoms of LDH [7]. Studies have concluded high prevalence of lumbar disc degeneration or LDH on magnetic resonance imaging (MRI) interpretations even in people without symptomatic LBP. These findings suggest that radiological parameters alone may not always align directly with clinical symptoms [8,9]. Thus, the above controversial findings from the evolving studies highlight the necessity to explore beyond the mechanical compression concepts. This includes immune dysregulation in pain and other symptoms underlying the pathophysiology of LDH.

Nucleus pulposus (NP) is being recognized as an immune-privileged organ not only due to its avascular nature but also due to sandwiched location within cartilaginous endplates. During the development from embryological notochord, NP is enclosed and forms a blood-NP barrier [10,11]. LDH leads to the disruption of immune privilege property of NP where the structural barrier between intervertebral disc (IVD) and immune system is disturbed, this unmasks the NP to the immune system and is increasingly recorded as a major event for LDH pathogenesis.

Increasing evidence targets the involvement of inflammation in generating pain and disease progression in LDH [11–15].

Studies have highlighted the persistent elevation of interleukin-6 (IL-6) and interleukin-8 (IL-8) with lumbar radicular pain[16,17]. Pro-inflammatory cytokines including IL-6, IL-8, interleukin 1 beta (IL-1β), and tumor necrotic factor alpha (TNF-α) are secreted by herniated disc itself and engaged in nociceptor sensitization, recruitment of immune cells, neuropathic pain and sciatica[7,11]. Despite these findings, the clinical utility of inflammatory markers in LDH remains incompletely understood. In addition, cellular functions depend on the programmed functions of actively transcribed genes, proteins and metabolites. Therefore, biological information in protein-protein interaction (PPI) network concept is vital for comprehensive understanding of biological systems. Most molecular functions that occur within a cell are organized by molecular machines constructed from a large number of proteins. Signaling within the proteins leads to dynamic PPIs, revealing the well-defined biological process, molecular functions and cellular components [18]. The present study aimed to quantify selected serum inflammatory cytokines, including interleukin-6 (IL-6), IL-8, tumor necrosis factor-alpha (TNF-α), and IL-1β, in patients with LDH and in age, gender, and body mass index (BMI) matched apparently healthy controls. The study further assessed the distribution and diagnostic ability of these cytokines in distinguishing patients from controls. In addition, correlations between cytokine levels and selected socio-demographic variables, clinical characteristics, and magnetic resonance imaging (MRI) findings were investigated in the LDH group. Furthermore, a PPI network was constructed to identify central hubs and functionally enriched pathways associated with LDH using computational analysis.

## Materials and Methods

### Study design

The present study employed a multi-faceted design to explore the role of circulating inflammatory cytokines such as IL-6, IL-1β, TNF-α and IL-8 in the LDH disease condition. As such, the study was compartmentalized into three well-defined study designs: A case-control design to assess the distribution of selected inflammatory cytokines between patients with LDH and healthy controls; a cross-sectional study to investigate the correlation of the above-mentioned inflammatory cytokines with clinical, socio-demographic and pre-operative lumbar MRI parameters, and a computational analysis to identify central hubs and functionally enriched pathways associated with the LDH disease context.

### Ethical approval

Approval was obtained from the Ethics Review Committee, Faculty of Medical Sciences, University of Sri Jayewardenepura, (Application reference number ERC 08/22). Separate information sheets were provided, and written informed consent were obtained individually from both patients with LDH and healthy controls prior to commencing the study.

### Sample size for case control study design

The sample size calculation formula, n = 2 (*Z_α_*_/2_ + *Z_β_*)^2^ (*σ*^2^/*d*^2^) [19] with *α* = 0.05 and *β* = 0.2 (80% power), according to a previous study, the population standard deviation for IL-6 was taken as 1.22pg/mL[20]. Present study expected to see a difference between means, that is, 60% the size of the standard deviation (*d* = 0.732 pg/mL). Thus, the minimum sample size calculated was 44 for each group. This case-control study was constructed to compare inflammatory cytokines distribution in patients with LDH (n=47) and age-gender and BMI-matched healthy controls (n=47).

### Study population

Patients (n=47), suffering from LBP and confirmed for LDH and undergoing lumbar microdiscectomy from selected hospitals in Sri Lanka and compared with age, gender and BMI matched equal number of apparently healthy controls (n=47) were recruited between 13^th^ December 2022 and 23^rd^ April 2024. Healthy controls had no previous history of any acute infections such as fever, chills, sore throat, or other infectious diseases for the past one-month duration, and had not consumed corticosteroids, non-steroidal anti-inflammatory drugs, or antibiotics during the above period. Further candidates diagnosed with acute or chronic inflammatory or infectious conditions persistent fever, cancer-associated cachexia, malignancies, autoimmune disorders, polyneuropathy and pregnant women were excluded from the study.

Patients enrolled for lumbar microdiscectomy were in the age range of 18-74 years and showed disc herniation extending from L1/L2 to L5/S1, they were not diagnosed for other spinal deformities such as congenital disc abnormalities, recurrent disc herniation, scoliosis, spondylosis or spondylolisthesis. An interviewer administered questionnaire was used to collect the baseline data including socio-demographic features, pre-operative lumbar MRI scan data and some clinical features such as lumbar radiating pain and walking difficulty were recorded.

### Quantification of selected circulating inflammatory cytokines

Blood specimens (3 mL) were collected from patients with LDH into a plain tube prior to surgery and before administration of anaesthesia. In addition, equal volume of blood specimens was also collected from matched healthy controls. Blood tubes were allowed to clot for 45 minutes at room temperature followed by centrifugation at 3000 rpm for 10 minutes to separate the serum. Further, serum was aliquoted into cryovials and frozen at −80°C for batch analysis. Serum inflammatory cytokines IL-6, IL-1β, IL-8 and TNF-α concentrations were quantified using enzyme-linked immunosorbent assay technique (Elabscience, Wuhan, China).

### Magnetic resonance imaging interpretations of lumbar spine

All patients with LDH underwent a regular T1 and T2 weighted pre-operative lumbar MRI scan prior to lumbar microdiscectomy. MRI details such as classification of disc herniation sub-types including protrusion, extrusion and sequestration, details regarding lumbosacral nerve root type and its compression information (L3 exit, L4 traverse, L4 exit, L5 exit, L5 traverse, and S1 traverse), details of cauda equina compression, occurrence of disc level herniation (single or multi-level herniation of lumbar discs) were documented.

### Statistical methods

Statistical assessments were carried out using Python (version 3.0). Baseline demographic variables, including age, BMI, were indicated as mean ± standard deviation and compared between groups using the independent samples t-test. Gender comparison was included as frequencies and percentages and compared using chi-square test. A p value < 0.05 was considered statistically significant. Mann Whitney U test was employed to determine the significant distribution of selected inflammatory cytokines between the two study populations (Patients with LDH and healthy controls). Further, ROC analysis was performed to assess the diagnostic ability of the cytokines as biomarkers. This included AUC, 95% confidence interval and optimal cut off of the markers to distinguish patients with LDH admitted for microdiscectomy over healthy controls.

Heatmap visualization was performed to investigate the complete distribution of the quantified cytokines (IL-1β, IL-6, IL-8, and TNF-α) across individual samples in the study group. Cytokine concentrations were z-score normalized across all the samples. Further, samples were categorized as control (0) and patients (1) to distinguish between patients and healthy controls. Cytokines were hierarchically clustered to investigate distinct cytokine distribution profiles with clear discriminative trends. Furthermore, Spearman’s correlation was performed for selected inflammatory cytokines against pre-operative lumbar MRI findings, socio-demographic data and clinical features recorded in patients with LDH and visualized using correlation heatmap annotated with correlation coefficients, and the corresponding p values were provided in S1 Fig under Supporting information.

### Construction of protein-protein interaction network for screening hub nodes and enrichment analysis data processing and fold change calculation

Group-level summary of cytokines from both patients with LDH and healthy controls were computed as means and standard error of the mean (SEM), fold change, Log₂-transformed fold change, statistical significance of differences of each cytokine between both the groups was calculated using Mann-Whitney U tests with p < 0.05 considered significant. Statistical analysis was done using Python version 3.0. The PPI network was constructed and analyzed using STRING version 12.0 and Cytoscape version 3.10.3. respectively.

### Construction of protein-protein interaction network

A dataset of cytokines and cytokine related proteins was downloaded from UniProt database [21] using https://www.uniprot.org/ with the (keyword:KW-0202) and reviewed (Swiss-Prot) filter, then accessed by selection of *Homo sapiens*. Altogether, 190 proteins were obtained from UniProt and input as seed proteins to the STRING database [22] with the confidence score 0.9 to extract strongly connected nodes (139) and edges (618) respectively and were imported into Cytoscape for downstream analysis.

Summary data from the experimental cytokines from both patients with LDH, and healthy controls were mapped onto this network as node attributes. Network topology was investigated using Network Analyzer. Molecular complex detection (MCODE) plugin [23] version 2.0.3 was accessed with the default parameters such as: Degree cutoff:2, Node score cutoff: 0.2, K-core:2, Max depth:100 to construct subnetworks with highly connected cluster of proteins from the entire PPI network. In addition, the above network was also further assessed to extract top ten highly connected hub nodes using maximal clique centrality (MCC) topological algorithm of Cytoscape cytoHubba plugin (version 0.1) [24]. Cytokines that were overlapping between the cytoHubba-derived top ten hub nodes and the highest-scoring MCODE subnetwork were chosen as input proteins for the investigation of functional enrichment pathways associated with the LDH context using the Database for Annotation, Visualization, and Integrated Discovery (DAVID) [25] https://david.ncifcrf.gov/ accessed on 26^th^ December 2025 including Gene Ontology terms (biological, and molecular), Kyoto encyclopedia of genes and genomes (KEGG) pathways. Enrichment significance was assessed using Fisher’s exact test. Multiple testing correction was done using the Benjamini–Hochberg false discovery rate method, and adjusted p-values < 0.05 were examined as statistically significant. PPI network analyses were executed at the protein level; however, gene symbols were used for standardized identification for database annotation, network construction, and downstream functional enrichment analyses.

## Results

### Study population and characteristics

This study included 94 participants, incorporating 47 patients with LDH and equal number of healthy controls (n = 47). The comparison of baseline features between both the study groups (Table 1) did not show any significant differences for mean age (p = 0.28), mean BMI (p = 0.53) and gender distribution (p = 0.67).

**Table 1.** Summary of demographic features comparison between patients with lumbar disc herniation and healthy controls.

|  | Patients with LDH | Healthy controls | p value | Total |
| --- | --- | --- | --- | --- |
|  | 47 | 47 |  | 94 |
| Male (n%) | 29 (61.7%) | 32 (68.09%) | 0.67 | 61 |
| Female (n%) | 18 (38.3%) | 15 (31.91%) |  | 33 |
| Age (Mean ± SD) years | 43.81 ± 15.56 | 40.26 ± 16.16 | 0.28 | |
| BMI (Mean ± SD) kgm<sup>-2</sup> | 26.32 ± 3.35 | 25.83 ± 4.21 | 0.53 | |

LDH, lumbar disc herniation; BMI, Body mass index

### Comparison of serum cytokine profiles in patients with lumbar disc herniation and healthy controls

As depicted in Table 2 serum IL-6 and IL-1β levels showed significant elevation in patients with LDH (n=47), compared to healthy controls (n=47) whereas TNF-α, IL-8 showed similar values in both groups.

**Table 2.**
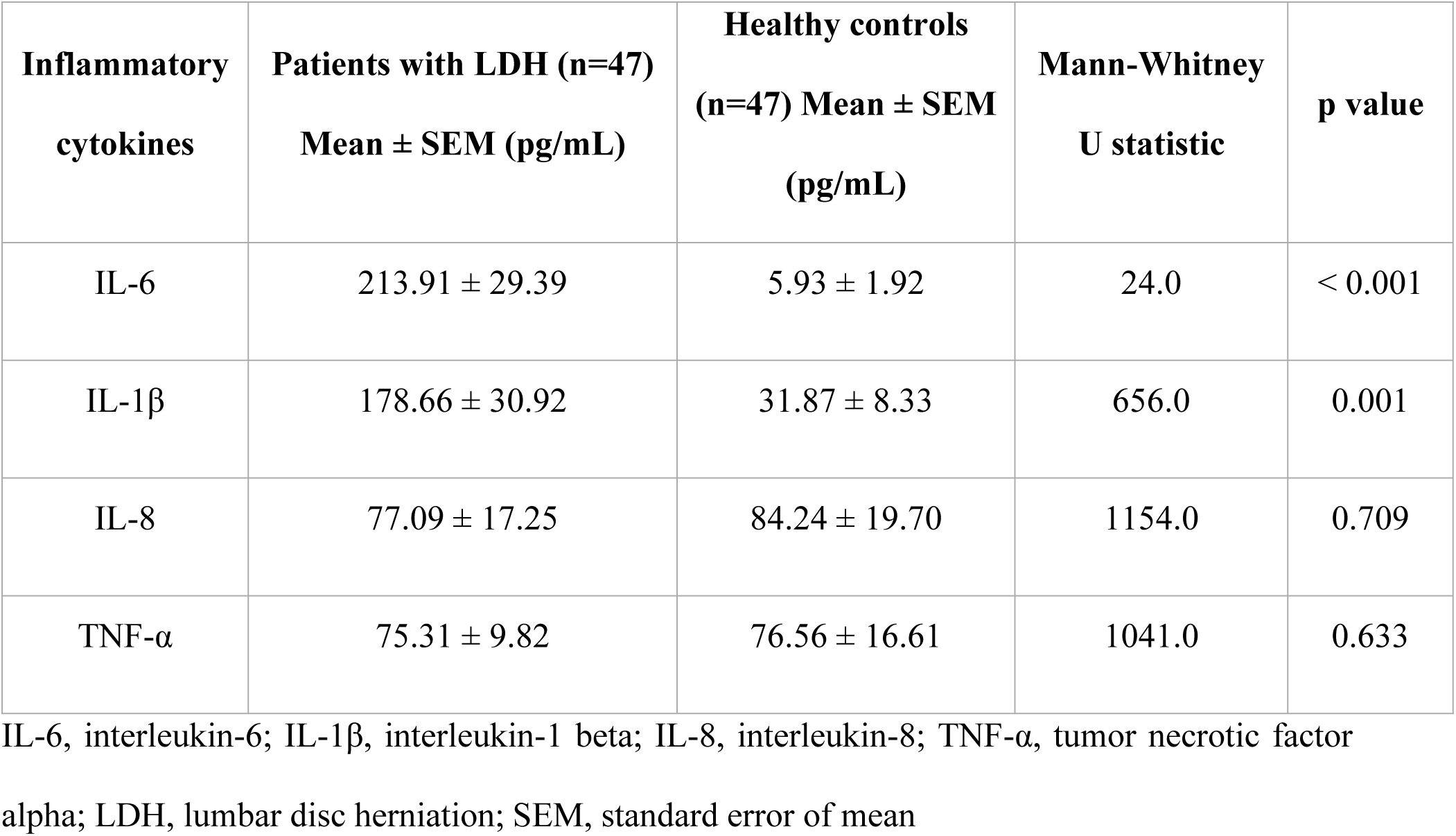
Serum inflammatory cytokines values in patients with lumbar disc herniation and healthy controls.

| Inflammatory cytokines | Patients with LDH (n=47)<br>Mean ± SEM (pg/mL) | Healthy controls<br>(n=47) Mean ± SEM<br>(pg/mL) | Mann-Whitney<br>U statistic | p value |
| --- | --- | --- | --- | --- |
| IL-6 | 213.91 ± 29.39 | 5.93 ± 1.92 | 24.0 | < 0.001 |
| IL-1β | 178.66 ± 30.92 | 31.87 ± 8.33 | 656.0 | 0.001 |
| IL-8 | 77.09 ± 17.25 | 84.24 ± 19.70 | 1154.0 | 0.709 |
| TNF-α | 75.31 ± 9.82 | 76.56 ± 16.61 | 1041.0 | 0.633 |

IL-6, interleukin-6; IL-1β , interleukin-1 beta; IL-8, interleukin-8; TNF-α , tumor necrotic factor alpha; LDH, lumbar disc herniation; SEM, standard error of mean

### Heatmap for the quantified cytokines data among patients with lumbar disc herniation and healthy controls

The heatmap revealed a colour based distribution of cytokine levels across individual samples in both the study groups as indicated in Fig 1.

**Fig 1.**
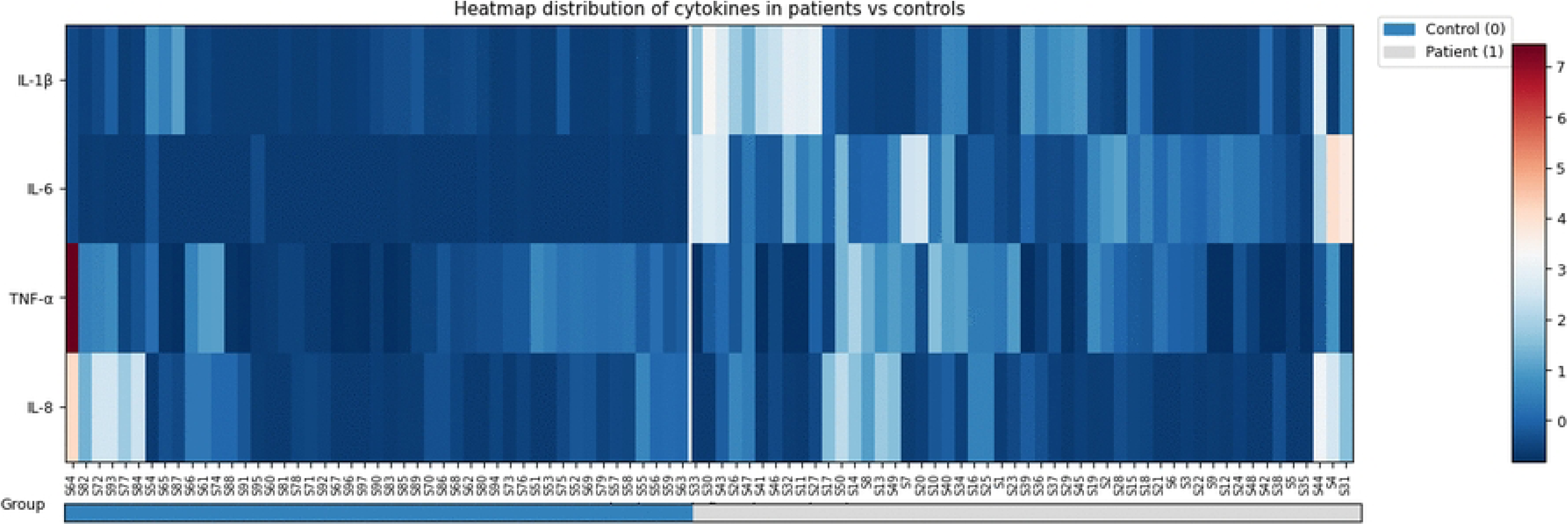
Heatmap of serum cytokine distribution in patients with lumbar disc herniation and healthy controls. Heatmap generated for the patients (n=47) and controls (n=47) depicts serum cytokine levels of every individual in columns and cytokines (IL-6, IL-8, TNF-α and IL-1β) in rows, where the cytokine quantified data representing healthy controls and patients, (Control (0) and (Patient (1)) indicated by the group bar below the heatmap. Color intensity reveals the variation in the z score normalized values of cytokines between both groups. IL-6, interleukin-6; IL-1β, interleukin-1 beta; TNF-α, tumor necrotic factor alpha; IL-8, interleukin-8

Accordingly, IL-6 and IL-1β showed elevation as well as diverging patterns of colour variability showing highly variable colour [varying from blue to dark red] and trending towards red shades among patients, and was uniformly lower and more homogenous in healthy controls group, indicating reduced cytokine quantification. Furthermore IL-8 and TNF-α exhibited uniformly low and less intense colours across both healthy controls and patients except for a small number of individuals displaying mild variations that didn’t show a discriminatory variation distribution among both the study groups.

### Receiver operating characteristic analysis to predict inflammatory cytokines as biomarkers

ROC curve analysis revealed predictive values for all the inflammatory cytokines and distinguishing patients with LDH and undergoing microdiscectomy from healthy controls as indicated in Table 3. IL-6 was found to be a more promising biomarker with greater AUC. The ROC curve analysis for IL-6 is shown in Fig 2.

**Fig 2.**
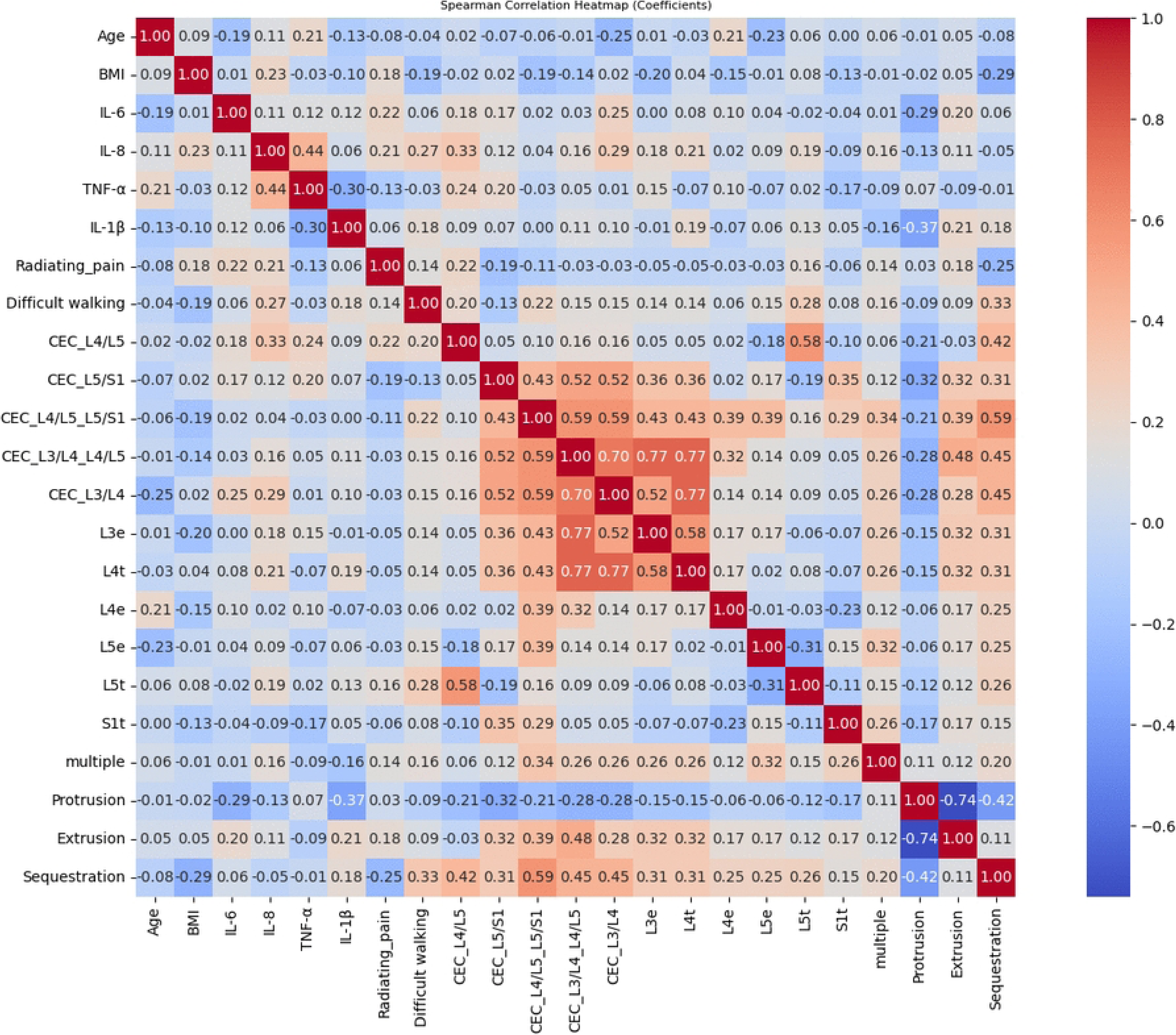
Receiver operating characteristic curve of serum interleukin-6 between study groups. Receiver operating characteristic curve displays the value for area under the curve (AUC) and the optimum cut off value for interleukin-6 (IL-6) to distinguish patients with lumbar disc herniation undergoing microdiscectomy from healthy controls

**Table 3.**
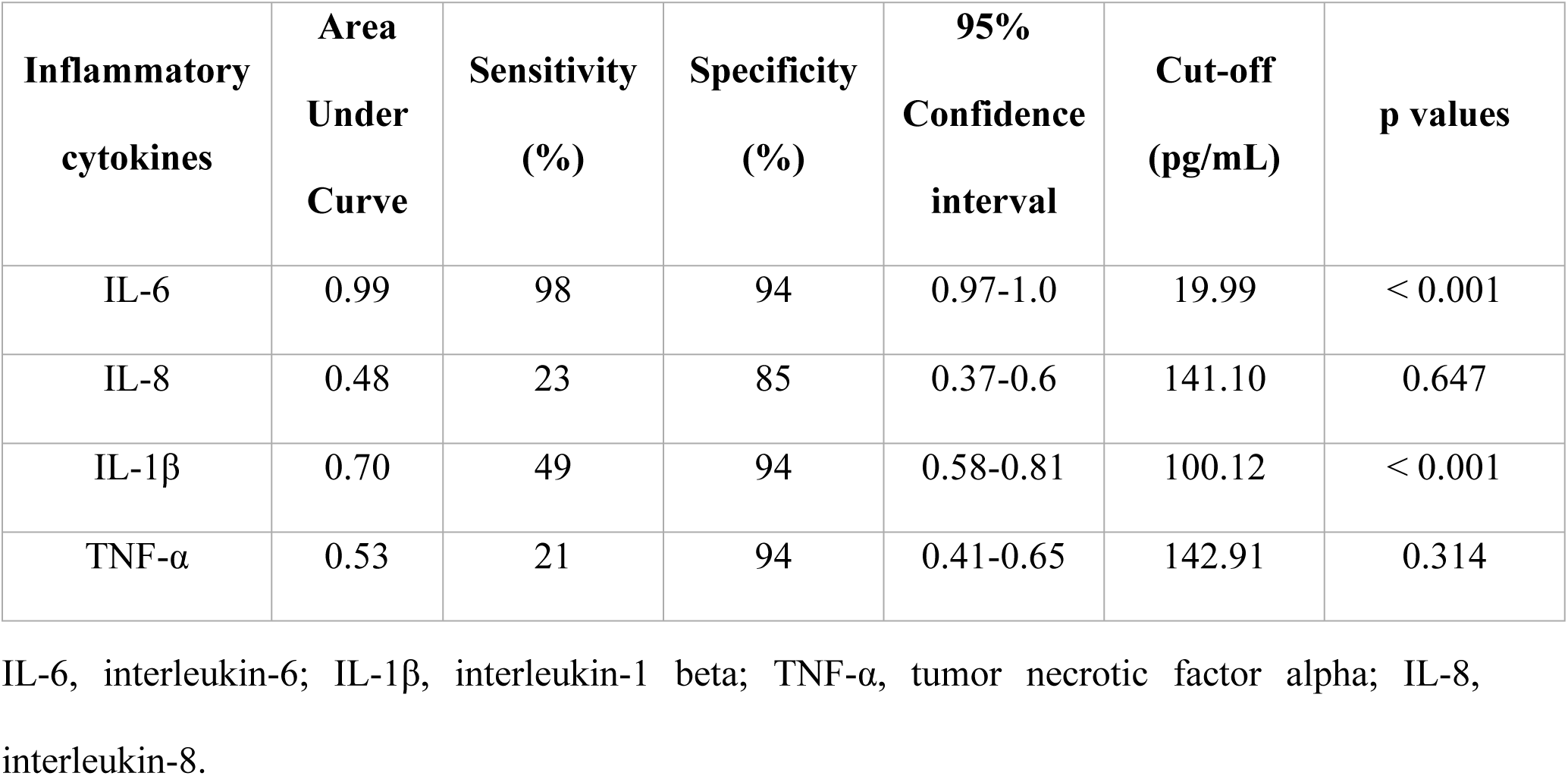
Receiver operating characteristic curve analysis for inflammatory cytokines.

### Correlation analysis between inflammatory cytokines and sociodemographic, clinical, and lumbar magnetic resonance imaging data

A Spearman correlation analysis resulted with positive and negative associations among the cytokines and other variables assessed as shown in Fig 3. while the corresponding p-values are indicated in S1 Fig under Supporting information.

**Fig 3.**
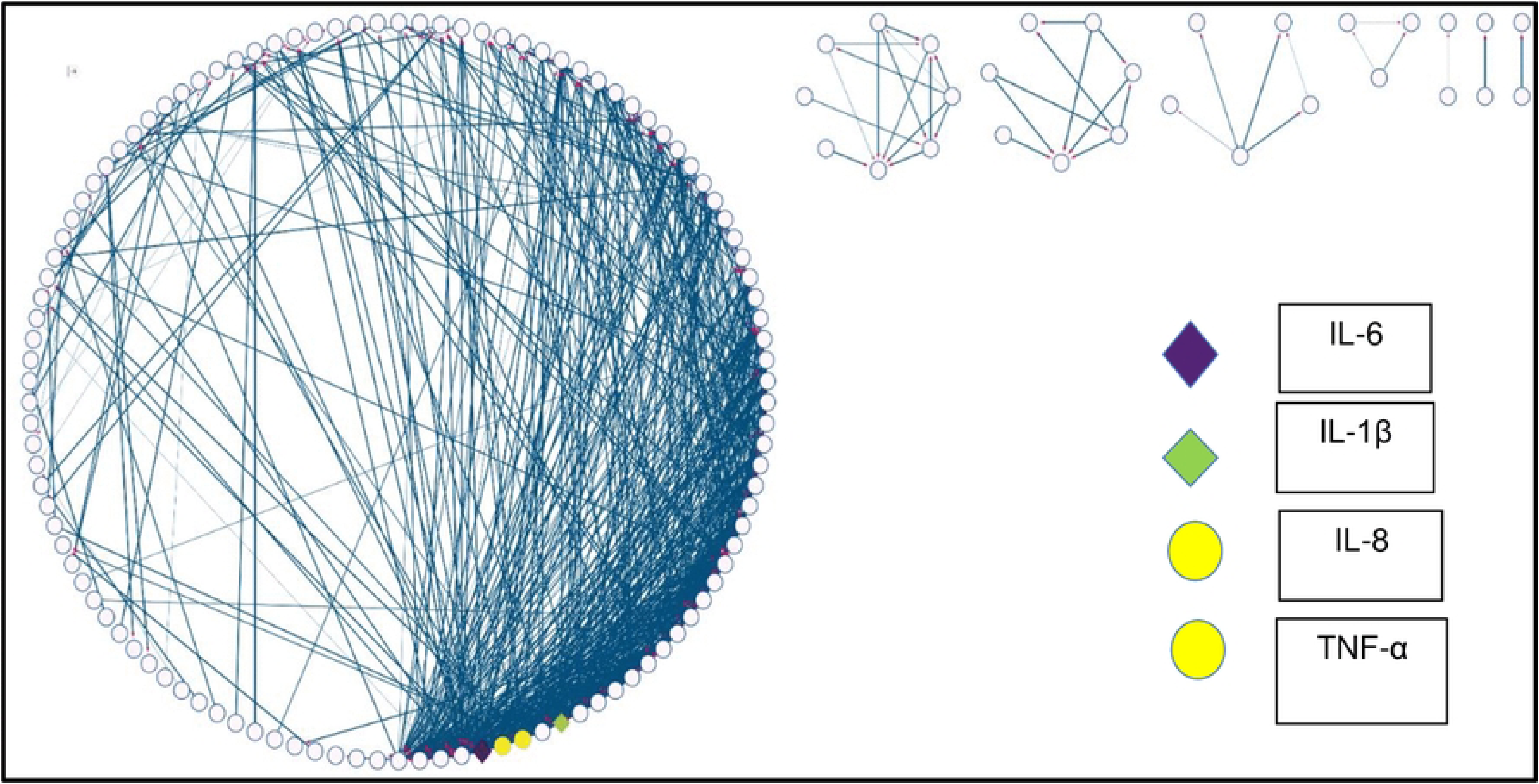
Correlation heatmap annotated with correlation coefficients among studied variables. IL-6, interleukin-6; IL-8, interleukin-8; TNF-α, tumor necrotic factor alpha; IL-1β, interleukin-1 beta; CEC_L4/L5, compression of cauda equina at L4/L5; CEC_L5/S1, compression of cauda equina at L5/S1; CEC_L4/L5_L5/S1, compression of cauda equina at both L4/L5 and L5/S1 disc levels; CEC_L3/L4_L4/L5, compression of cauda equina at both L3/L4 and L4/L5 disc levels; CEC_L3/L4, compression of cauda equina at L3/L4; L3e, compression in L3 exit nerve root; L4t, compression in L4 traverse nerve root; L4e, compression in L4 exit nerve root; L5e, compression in L5 exiting nerve root; L5t, compression in L5 traversing nerve root; S1t, compression in S1 traversing nerve root; multiple, multiple disc herniation

#### Correlation analysis of inflammatory cytokines and clinical variables

A Spearman correlation analysis identified significant relationships between cytokine levels and selected pre-operative lumbar MRI, socio-demographic and clinical features as summarized below. Inflammatory cytokines IL-8 and TNF-α exhibited a significant moderate positive correlation (r = 0.44, p = 0.001), while IL-1β and TNF-α levels showed significant negative correlation (r = −0.3, p = 0.036). Further, IL-1β showed a significant negative correlation with protrusion disc herniation subtype (r = −0.37, p = 0.008), where extrusion (r = 0.21, p = 0.14) and sequestration (r = 0.18, p = 0.204) resulted with non-significant positive correlation. Moreover, serum IL-8 levels and difficulty in walking (r = 0.27, p = 0.054) showed positive correlation, although the correlation was not statistically significant.

#### Disc herniation subtypes and walking difficulty

Sequestration demonstrated a positive association with difficulty in walking (r = 0.33, p = 0.019), while protrusion indicated a negative trend (r = −0.09, p = 0.538).

#### Involvement of lumbosacral nerve roots with subtypes of disc herniation

Analysis of lumbosacral nerve root involvement revealed that extrusion and sequestration disc herniation subtypes were positively correlated with the compression of multiple nerve roots, including L3 exit, L4 exit, L4 traversing, L5 exit, L5 traversing, and S1 traversing roots, whereas protrusion showed a persistent negative correlation.

#### Cauda equina compression and associated parameters

Among the analyzed cytokines, IL-8 showed a significant positive correlation with cauda equina compression at single disc L4/L5 (r = 0.33, p = 0.019) and L3/L4 (r = 0.29, p = 0.038) levels. In contrast, no significant association was reported between IL-8 and multilevel cauda equina compression. In addition, L5 traverse nerve root involvement showed significant positive correlation with the compression of cauda equina at the disc level L4/L5 (r = 0.58, p < 0.001). Sequestration demonstrated significant positive correlations with multilevel cauda equina compression. Sequestration with multiple disc levels L4/L5 with L5/S1 (r = 0.59, p < 0.001) and L3/L4 with L4/L5 (r = 0.45, p = 0.001) was observed. Similarly, extrusion also showed significant positive correlations in the presence of cauda equina compression at these respective disc levels L4/L5 with L5/S1 (r = 0.39, p = 0.005), and L3/L4 with L4/L5 (r = 0.48, p < 0.001). Whereas, protrusion exhibited a significant negative correlation with compression of cauda equina at the same respective disc levels L4/L5 with L5/S1(r = –0.21, p = 0.142) and L3/L4 with L4/L5 (r = −0.28, p = 0.038).

#### Association of radiating pain with cytokine levels and neural compression

Significant correlations were not reported between radiating pain and cytokine levels or different levels of compression with cauda equina and nerve root involvement (p > 0.05).

### Identification of hub proteins from protein-protein interaction network

Topological network assessment revealed experimentally assessed cytokines (IL-6, TNF-α, IL-8, and IL-1β) align with highly interconnected regions, as illustrated in Fig 4. In addition, MCODE analysis revealed the presence of aforementioned cytokines within the prominent, highest-scoring inflammatory subnetwork, indicating a densely interacting subnetwork is associated with inflammatory responses as tabulated in Table 4. Furthermore, combination of experimental cytokine data with the network confirmed that IL-1β, IL-8, IL-6, and TNF-α ranked within the top four nodes according to the MCC centrality score from cytoHubba Cytoscape plugin. This indicates their significant prominence as hubs in the cytokine cascade as shown in Fig 5. Overlapping cytokines from MCODE and cytoHubba analysis identified IL-8, TNF-α, IL-6, IL-1β, IL-1α, CSF2, CSF3, and IL-10 as central hubs.

**Fig 4.**
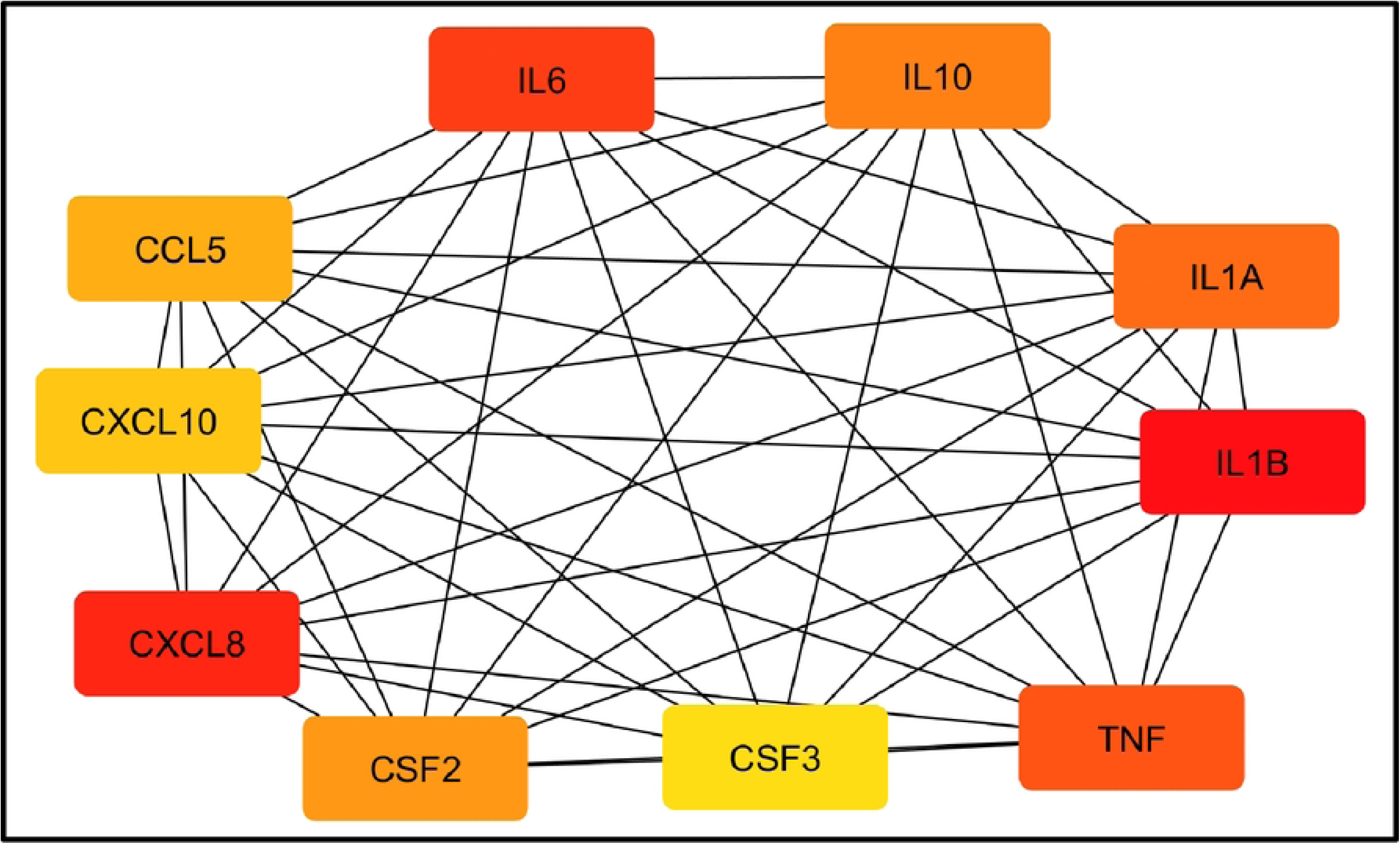
Protein-protein interaction network of high-confidence human cytokines in lumbar disc herniation. In protein–protein interaction network experimentally quantified cytokines are indicated as colored nodes. Colorless nodes indicate the rest of the human cytokines that are interconnected with 0.9 confidence score. Node color varies with the fold change of cytokines in patients with lumbar disc herniation compared with controls, ranging in an ascending order of the fold change from yellow to green to violet. Diamond-shaped nodes indicate significantly upregulated cytokines, while rest of the colored ellipse-shaped nodes represent non-significant cytokines. The interaction network was constructed using Cytoscape, where degree sorted circle layout was employed. Blue colored lines between the nodes (edges) portray active interactions or protein-protein interactions.IL-6, interleukin-6; IL-1β, interleukin-1 beta; IL-8, interleukin-8; TNF-α, tumour necrotic factor alpha

**Fig 5.**
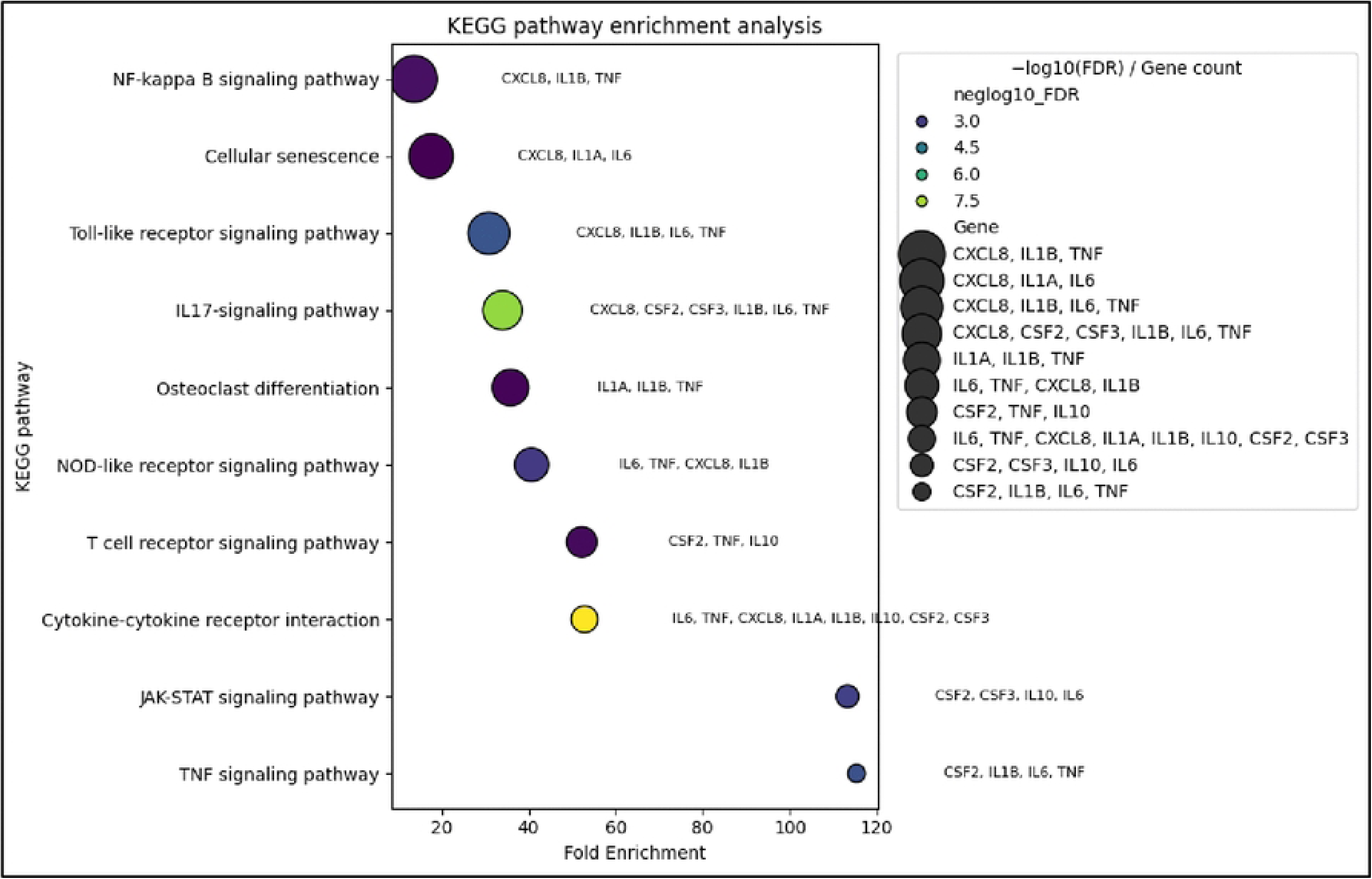
Maximal clique centrality ranked top ten hub nodes in the protein–protein interaction network. Protein-protein interaction network hubs are coloured in accordance with maximal clique centrality (MCC) scores received from cytoHubba assessment. Nodes with low MCC scores are indicated in yellow, moderate MCC scores in orange, and the highest MCC-score hubs are in red. Gene symbols are represented as standardized identifiers corresponding to cytokine and cytokine-related proteins. TNF, Tumor necrotic factor alpha; CXCL10, C-X-C motif chemokine 10; IL1B, interleukin 1 beta; IL1A, interleukin 1 alpha; IL10, interleukin 10; CCL5, C-C motif chemokine 5; CXCL8, interleukin 8; IL6, interleukin 6; CSF2, colony-stimulating factor 2; CSF3, colony-stimulating factor 3.

**Table 4.** Summary of subnetworks equal or beyond the cluster score 5.0 retrieved from the protein-protein interaction network using molecular complex detection (MCODE) algorithm including cluster scores, number of nodes, and number of edges and nodes details.

| Cluster No | Cluster score | No. of nodes | No. of edges | Nodes |
| --- | --- | --- | --- | --- |
| 1 | 13.33 | 28 | 180 | CCL3, CCL2, XCL1, IL13, IL6, IL1B, CXCL14, CCL26, CCL13, CXCL8, CSF2, CXCL1, PPBP, IL1A, TNF, IFNG, PF4, CXCL12, CCL21, IL2, IL10, IL17A, PF4V1, CXCL9, CSF3, IL4, CCL4, IL5 |
| 2 | 6.364 | 12 | 35 | CCL20, CXCL2, CXCL11, CCL19, CXCL10, CCL25, CCL17, CX3CL1, CXCL3, CCL22, CCL5, CCL11 |
| 3 | 5.0 | 5 | 10 | BMP2, BMP7, BMP4, GREM2, GREM1 |

Gene symbols show the cytokine or cytokine-related proteins retrieved from the interaction network.

CCL3,C-C motif chemokine 3; CCL2, C-C motif chemokine 2; XCL1, lymphotactin; IL13, interleukin 13; IL6, interleukin 6; IL1B, interleukin 1 beta; CXCL14, C-X-C motif chemokine 14; CCL26, C-C motif chemokine 26; CCL13, C-C motif chemokine 13; CXCL8, interleukin 8; CSF2, colony stimulating factor 2; CXCL1, C-X-C motif chemokine 1; PPBP, Platelet basic protein; IL1A, interleukin 1 alpha; TNF, tumor necrotic factor alpha; IFNG, interferon gamma; PF4, platelet factor 4; CXCL12, C-X-C motif chemokine 12; CCL21, C-C motif chemokine 21; IL2, interleukin 2; IL10, interleukin 10; IL17A, interleukin 17A; PF4V1, platelet factor 4 variant; CXCL9, C-X-C motif chemokine 9; CSF3, colony stimulating factor 3; IL4, interleukin 4; CCL4, C-C motif chemokine 4; IL5, interleukin 5; CCL20, C-C motif chemokine 20; CXCL2, C-X-C motif chemokine 2; CXCL11, C-X-C motif chemokine 11; CCL19, C-C motif chemokine 19; CXCL10, C-X-C motif chemokine 10; CCL25, C-C motif chemokine 25; CCL17, C-C motif chemokine 17; CX3CL1, C-X3-C motif chemokine 1; CXCL3, C-X-C motif chemokine 3; CCL22, C-C motif chemokine 22; CCL5, C-C motif chemokine 5; CCL11, C-C motif chemokine 11; BMP2, bone morphogenetic protein 2; BMP7, bone morphogenetic protein 7; BMP4; bone morphogenetic protein 4; GREM1, gremlin-1; GREM2, gremlin-2.

### Functional enrichment analysis

#### Kyoto encyclopedia of genes and genomes (KEGG) pathways

DAVID-based functional enrichment analysis demonstrated significant enrichment of inflammatory, apoptotic, and cytokine signaling pathways. KEGG pathway annotation detected multiple signaling pathways demonstrated by the enriched genes including cytokine-cytokine receptor interaction, interleukin 17 (IL-17) signaling pathway, Toll-like receptor (TLR) signaling pathway, TNF signaling pathway, Janus kinase-signal transduction and activation of transcription (JAK-STAT) signaling pathway, NOD-like receptor (NLR) signaling pathway, Nuclear factor-kappa B (NF-ĸB) signaling pathway, T cell receptor signaling pathway, cellular senescence, and osteoclast differentiation as illustrated in Fig 6.

**Fig 6.**
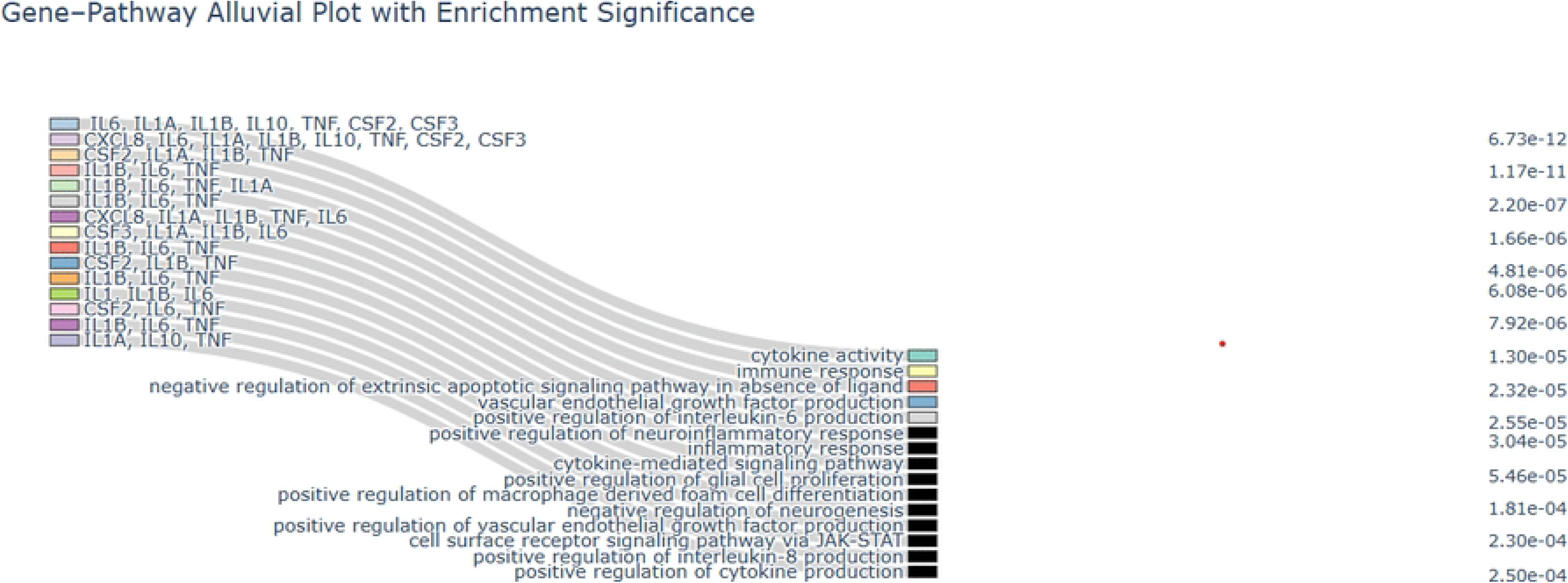
Dot plot shows significantly enriched Kyoto encyclopedia of genes and genomes (KEGG) pathways. In the dot plot, fold enrichment is assembled on the x-axis, while KEGG pathways are listed on the y-axis. Dot color shows the pathway represented as −log10 Benjamini–Hochberg adjusted p-values. The cytokine genes contributing to each pathway are annotated adjacent to the corresponding dots. NF-ĸB, Nuclear factor kappa B; JAK-STAT, Janus kinase-signal transduction and activation of transcription; TNF, tumor necrotic factor alpha; CXCL10, C-X-C motif chemokine 10; IL1B, interleukin 1 beta; IL1A, interleukin 1 alpha; IL10, interleukin 10; CCL5, C-C motif chemokine 5; CXCL1, growth regulated alpha protein; CXCL8, interleukin 8; IL6, interleukin 6; CSF2, colony-stimulating factor 2; CSF3, colony-stimulating factor 3.

#### Gene Ontology

The top ten modulated gene ontology biological processes, and molecular processes with enriched proteins combination is illustrated in Fig 7.

**Fig 7.**
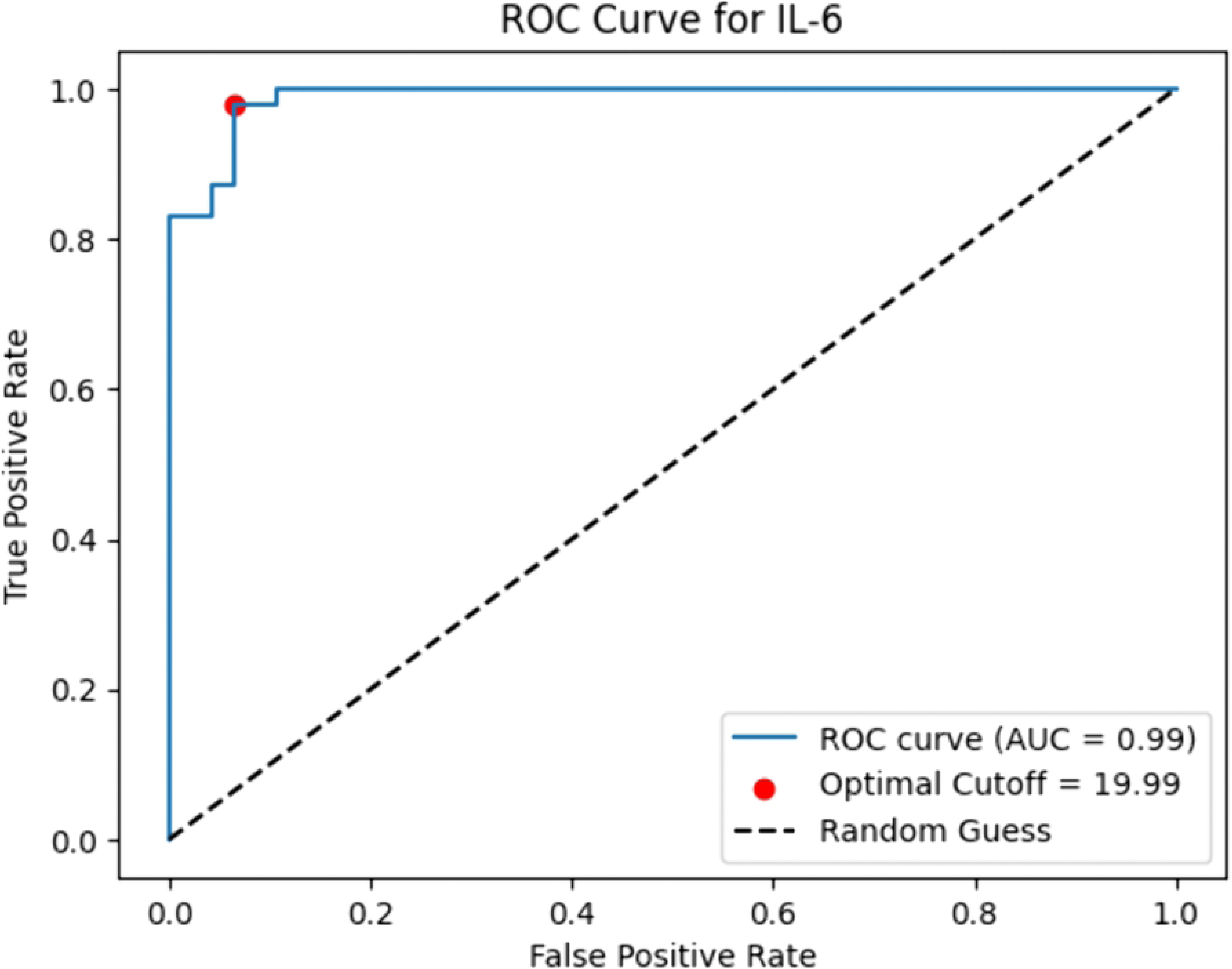
Gene-Pathway Alluvial plot with significantly enriched Gene Ontology terms (Biological and Molecular) JAK-STAT, Janus kinase-signal transduction and activation of transcription; IL6, interleukin 6; TNF, tumor necrotic factor alpha; CXCL8, interleukin 8; IL1A, interleukin 1 alpha; IL1B, interleukin 1 beta; IL10, interleukin 10; CSF2, colony stimulating factor 2; CSF3, colony stimulating factor 3;

## Discussion

This study presents a comprehensive integrative approach to the discovery of biomarker in LDH by combining experimental cytokine quantification with diagnostic evaluation, correlation with multiple variables such as socio demographic, radiological and clinical features associated with patients and integration of experimental work with the in-silico network and functional enrichment pathways. Unlike previous studies that have primarily focused on individual aspects of inflammation, the present work incorporates multiple experimental layers to explore a system level understanding of the involvement of cytokine in LDH. The integration of quantitative cytokine data with PPI networks and pathway enrichment analysis offers additional insight into the interconnected biological mechanisms underlying LDH disease pathology.

### Receiver operative characteristic analysis for inflammatory cytokines in patients with lumbar disc herniation and healthy controls

This study showed significant elevation in the distribution of serum IL-6 and IL-1β in patients with LDH compared to age, gender and BMI matched healthy controls. The above findings provide evidence that inflammatory cytokines are significantly associated with the pathophysiology of LDH.-Coinciding with previous findings highlighting IL-6 as a key pro-inflammatory cytokine involving amplified inflammatory reactions, local pain and systemic inflammation in the context of LDH [26,27]. The increased concentration of IL-6 in patients with LDH has been documented in similar studies [20,28,29], highlighting its role as a selective biomarker. Importantly, the ROC analysis resulted with IL-6 as an optimistic biomarker for segregating patients with LDH from healthy individuals, with an AUC of 0.99, indicating a promising diagnostic performance. The calculated cut-off value of 19.99 pg/mL for IL-6 derived high sensitivity (99%) and specificity (94%), emphasizing its reliability as a diagnostic indicator. To our knowledge, this is the first study that demonstrates IL-6 as a promising biomarker to segregate patients with LDH undergoing surgical intervention from healthy controls. Future studies with larger study population and longitudinal follow-up are recommended to confirm these biomarkers applicability in clinical practice.

### Inflammatory cytokine correlation with radiological, clinical and demographic data

The present study also investigated the inter-correlation with the inflammatory cytokine levels, and revealed inflammatory cross talks between structural anatomy of herniated discs of lumbar spine, clinical features as well as socio-demographic data.

A moderate positive correlation between IL-8 and TNF-α (r = 0.44, p = 0.001) suggests a combined engagement and regulated inflammatory pathway involving these both inflammatory cytokines, which may impact the chronic neuroinflammation in LDH concept. TNF-α is a multi-functional pro-inflammatory cytokine that plays a major role in regulating immune responses, apoptosis and inflammation in the disease context. Further, TNF-α stimulates the production of IL-8 with the relevant activated nuclear factor kappa B (NF-ĸB) pathway and phosphoinositide 3-kinase (PI3K/Akt) pathway [30]. Conversely, IL-1β showed a significant negative correlation with TNF-α (r = −0.3, p = 0.036) indicating a robust regulatory mechanism within the inflammatory pathway. Importantly, our study demonstrated a significant variation of IL-1β levels across disc herniation subtypes where IL-1β demonstrated a significant negative correlation with protrusion (r = −0.37, p = 0.008), mild positive associations with extrusion (r = 0.21, p = 0.140) and sequestration (r=0.18, p=0.204). These findings indicate a positive trend towards IL-1β levels in non-contained disc herniation subtype.

The above concept can be further explained using a robust connection applied via the arachidonic acid cascade. Previously reported study has confirmed the presence of higher levels of Leukotriene B4 (LTB4) and thromboxane B2 biomarkers in non-contained disc herniations compared to contained herniation type [31]. While literature particularly connecting IL-1β expression levels to the distinct stages was not interpreted, studies have revealed a clear association between disc herniation subtypes and elevated levels of LTB4. This suggests a probable mechanism; the contained nature and the morphology of AF trigger a localized inflammatory response, leading to the synthesis of LTB4. Further, LTB4, in turn, amplifies the inflammatory cascade by inducing the production of IL-1β [32,33] by increasing the activation of NF-ĸB Pathway [34]. Therefore, in our study a positive trend in IL-1β expression with the containment disc concept is likely due to LTB4-mediated inflammatory response. Thus, the above concept underscores a critical positive regulation, where both IL-1β and LTB4 link to the persistent inflammation and degenerative processes observed in disc pathology. Thus, the above concept suggests that IL-1β may function as a biomarker to screen disc herniation subtype as contained and non-contained disc herniation. The significant association between IL-8 and single-level cauda equina compression suggests the contribution of IL-8 in specific disc pathology. IL-8 is known to play an important role in neutrophil recruitment and neuroinflammatory signaling. In contrast, the absence of a significant association with multilevel cauda equina compression may be attributed to lesser number of patients reported with multilevel cauda equina compression in this study, reducing statistical power or may engage in a more complex inflammatory feedback loop.

Further, IL-8 and its borderline association with walking difficulty (r = 0.27, p = 0.054) indicates a potential role of IL-8 in modulating pain and mobility-related outcomes, while the correlation with walking difficulty did not reach statistical significance, the observed trend supports the inflammation related to walking difficulty in patients with LDH. The above finding is further supported by a study by Tian *et al*. [35] confirmed that the human IL-8 (hIL-8) transgenic mice that express IL-8 in the IVD and articular cartilage, travelled shorter distances and spent significantly minimal time in locomotion compared to their control littermates, suggesting that hIL-8 influences their behavioural patterns.

Among herniation subtypes, sequestration showed a significant positive correlation with walking difficulty (r = 0.33, p = 0.019). Sequestration and functional impairment like difficulty in walking underscores the clinical influence of disc pathology. Further, the above results support the close connection between the type of structural disruption and functional limitation. MRI is restricted in analyzing free fragment disc between the lumbar nerve roots and dural theca, and dorsal to the dural sac. Nevertheless, detached herniated disc’s size, composition, and location determine the different outcomes [36].

Among disc herniation subtypes, sequestration showed significant positive correlation with cauda equina compression across multiple disc levels L4/L5 with L5/S1 (r = 0.59, p < 0.001) and L3/L4 with L4/L5 (r = 0.45, p = 0.001), whereas, protrusion exhibited a negative correlation with cauda equina compression at the same respective disc levels L4/L5 with L5/S1 (r = –0.21, p = 0.142) and L3/L4 with L4/L5 (r = −0.28, p = 0.048). The negative and the positive correlation obtained for disc herniation subtypes with the compression of cauda equina reveals that, disc protrusion maintains the structural containment within the AF, minimizing the exposure of disc material towards the compression of cauda equina. The anatomical variation behind the structural disruption of disc herniation subtypes may therefore impact the intensity in the compression of cauda equina and influencing their role as major mediators in disease background. The above results reveal a robust anatomical consistency observed between the compression of cauda equina with disc herniation subtype, where central disc orientation dominates and further, findings validate the combination of anatomical inference with radiological interpretations. Although multilevel cauda equina compression reported significant associations with sequestration subtypes, the very small number of patients restrict the strength of the above findings. Therefore, further experiments are warranted in larger cohorts.

Overall, the results emphasized multiple inflammatory pathways and lumbosacral nerve root involvement across disc herniation subtypes, contributing to the concept that cytokine quantification in conjunction with lumbar spine MRI findings may lead to a more detailed framework for understanding the theory about the variability in LDH.

### Bioinformatics analysis on protein-protein interaction network construction

Network-based system biology reveals an understanding about the proteins interactions in explaining the disease pathophysiology [37]. Network analysis revealed IL-1β as the top hub node ranking with the highest MCC score from cytoHubba analysis, followed by IL-8, IL-6, and TNF-α etc. Hub proteins discovered from the study, may function as reliable molecules for additional functional validation studies and greatly influence LDH pathophysiology. Their position as central nodes reflect their multi- regulatory mechanisms mainly on inflammatory cascades of reactions relevant to disc pathology and neuroimmune interactions associated with disc pathology. Functional enrichment analysis using the DAVID database showed the major influence of cytokine storm associated pathways, predominantly cytokine-mediated inflammatory cascades play an important part of inter-cellular crosstalk in the context of LDH.

The above findings remain concordant with the signaling of multiple inflammatory pathways such as enrichment of IL-17, TNF-α, and NF-ĸB signaling pathways. Further, TLR plays a major role in the activation of glial cells and neurons distributed in the spinal cord and might impact nociceptive sensitization with the nerve injury [38]. NOD-like receptor (NLR) signaling pathways, suggest an active immune response. Activation of NLRP3 and caspase-1 have also been explained in degenerated discs that lead to inflammasome activation and the secretion of IL-1β leads to disc degeneration [39].

Further the activation of NF-ĸB signaling leads to the enrichment supports the exponential escalation of pro-inflammatory gene expression of IL-1α, IL-1β, IL-6, IL-8, IL-17 and TNF-α with the disc degeneration [40]. The JAK-STAT signaling pathway shows the involvement of moderate number of cytokines, and cytokine-mediated signal transduction, which complements the observed cytokine-cytokine receptor interactions. This pathway is important in the transcription of genes responsible for immune responses, cell proliferation, and differentiation [41]. IL17A positively transcripts the activation of JAK-STAT pathway by inducing the rapid tyrosine phosphorylation of the JAK family and STAT 1,2,3, and 4 [42], further, it influences in the upregulation of vascular endothelial growth factor (VEGF) through JAK-STAT pathway, often the elevated levels of IL17A and VEGF could be observed in degenerated discs [43], T cell receptor signaling pathway describes the adaptive immune system influence and suggesting a true interconnection between innate and adaptive immunity including T cell activation and differentiation; often indicating chronic and persistent inflammation. Osteoclasts play a major role in triggering LBP, by inducing porosity of endplates with sensory innervation into the porous regions [44,45]. Osteoclasts often show characteristic features of senescence during osteoclastogenesis [46]. Cellular senescence is a crucial trigger for disc degeneration through the senescence-associated secretory phenotype by secreting pro-inflammatory cytokines, matrix degrading proteases, growth factors, angiogenic factors and chemokines that leads to immune dysregulation and worsen the degenerative symptoms via autocrine and paracrine pathways in the context of LDH [47–49]. Cytokines such as IL-6, TNF-α, IL-8, IL-1α, IL-1β, IL-10, CSF2, and CSF3, represented as central hubs with pleiotropic effects. The occurrence of both pro-inflammatory cytokines (IL-1β, IL-6, TNF-α, IL-8) and the anti-inflammatory cytokine IL-10 deals with a complex regulatory environment where inflammatory activation is regulated to maintain homeostasis in the inflammatory micro-environment of the herniated disc. In addition, exploring the balance maintained between pro-inflammatory and anti-inflammatory signals are important to develop therapeutic strategies in the concept of LDH disease. The above scenario represents a mechanistic overlap with inflammation. Together, these findings not only validate the major inflammatory cytokines measured experimentally but also investigate an expanded molecular network and signaling architecture that may drive neuro-inflammation and nerve sensitization in LDH.

## Conclusions

The results confirmed a significant elevation of IL-6 and IL-1β in patients with LDH, with IL-6 indicating a remarkable diagnostic performance contributing a major role as a predictive biomarker. Further, IL-1β demonstrated its ability to serve as a biomarker in screening contained and non-contained disc herniation subtype. In addition, IL-8 showed significant positive correlations with cauda equina compression occurred at single disc levels L4/L5 and L3/L4. This is the first study to assess cytokine correlations with the involvement of cauda equina compression. In silico bioinformatics analysis identified central inflammatory mediators including IL-1β IL-6, IL-8, TNF-α, CSF2, CSF3, IL-1α, and IL-10, with IL-1β emerging as the major hub. Functional enrichment analysis using the DAVID database consistently identified pathways related to inflammatory response, apoptotic regulation, and cytokine signaling across both analyses. These findings strengthen the central role of the key hubs in mediating interconnected inflammatory and regulatory processes in LDH. Future studies focusing on targeted anti-inflammatory therapeutic strategies may open new avenues for personalized treatment options in the concept of LDH.

## Data Availability

All relevant data are within the manuscript and its Supporting Information files. Additional data underlying the findings of this study are available from the corresponding author upon reasonable request with limitations, however no individual identifiers will be provided. Data cannot be made publicly available due to ethical restrictions involving human participants, as imposed by the Ethics Review Committee of the University of Sri Jayewardenepura.

## Acknowledgments

We would like to thank Ms. Sriyani Hewaratne, Technical Officer, Department of Medicine, University of Sri Jayewardenepura for technical assistance

## Supporting information

**S1 Fig. Heatmap illustrating p-values of Spearman correlation analysis among cytokines, socio-demographic, clinical parameters, and MRI features.**

The heatmap shows the statistical significance (p-values) of Spearman correlations between inflammatory cytokines (IL-6, IL-8, TNF-α, and IL-1β), demographic variables (age and BMI), clinical parameters (radiating pain and difficulty walking), and lumbar MRI parameters. Each cell represents the p-value corresponding to the association between two variables, with color intensity depicting the level of statistical significance.

## Notes

### Competing Interest Statement

The authors have declared no competing interest.

### Funding Statement

Yes

### Author Declarations

Ethical approval was obtained Ethics review committee of Faculty of Medical Sciences, University of Sri Jayewardenepura and written informed consent was obtained. Patient details will be maintained at high confidentiality, etc

